# Family income is associated with regional brain glucose metabolism in middle-aged adults

**DOI:** 10.1101/2024.09.18.24313859

**Authors:** Kyoungjune Pak, Seunghyeon Shin, Hyun-Yeol Nam, Keunyoung Kim, Jihyun Kim, Myung Jun Lee

**Author notes:** Kyoungjune Pak (Corresponding author) Dept. of Nuclear Medicine, Pusan National University Hospital, 179 Gudeok-ro, Seo-gu, Busan 49241, Republic of Korea, Seunghyeon Shin (Corresponding author). Kyoungjune Pak and Seunghyeon Shin contributed equally in this work.

## Abstract

Socioeconomic status is a multifaceted construct that plays a prominent role in shaping our environment by defining our access to healthcare, nutrition, and enrichment, as well as represents social standing. Therefore, to address the effects of family income, and education level on brain glucose metabolism, we analyzed a large cohort of healthy middle-aged adults who underwent brain ^18^F-FDG PET, and survey of family income and education level. We retrospectively analyzed data of healthy males who underwent health check-up program. Health check-up program included 1) Brain ^18^F-FDG PET, 2) anthropometric measurements, 3) survey of family income and education level, and 4) measures of stress, anxiety, and depression. The effects of family income and education level on regional SUVR were investigated using Bayesian hierarchical modelling. A total of 233 healthy males were included in this study. Family income was positively correlated with education level. There was no significant indirect effect of family income or education level via stress, anxiety, or depression on regional brain glucose metabolism. Family income is positively associated with brain glucose metabolism in caudate, putamen, anterior cingulate, hippocampus, and amygdala, while education level does not show any significant association with brain glucose metabolism in middle-aged adults. In conclusion, family income is positively associated with brain glucose metabolism in caudate, putamen, anterior cingulate, hippocampus, and amygdala, while education level does not show any significant association with brain glucose metabolism. This finding might reflect the link between family income, and reward sensitivity, stress in middle-aged adults.

## INTRODUCTION

Brain is a social organ ^1^. No human brain exists outside of a particular socioeconomic context ^2^. Socioeconomic status (SES) is a multifaceted construct that plays a prominent role in shaping our environment by defining our access to healthcare, nutrition, and enrichment, as well as represents social standing ^3^. SES is predictive of a broad range of important life outcomes ^2^. Higher SES is associated with better health both physically, and mentally. The incidence of heart disease, stroke, cancer, diabetes, as well as depression, anxiety, and psychosis is positively related with SES ^4^. Also, intelligence and academic achievement exhibit positive gradients with SES ^2^.

Even after 3 months of social housing, dopamine receptor (DR) availability was increased in dominant monkeys, compared with subordinate monkeys or the baseline state of the individual housing ^5^. In addition, subordinate monkeys were found to be vulnerable to the reinforcing effects of cocaine, while dominant monkeys were found to be resistant to its effects ^5^. Therefore, these neurobiological alterations in DR availability by social context can lead to different behavioral phenotypes ^5^.

Examination of the relationship between SES, and the brain has primarily focused on the earlier or later stages of the lifespan when the brain is most vulnerable, which are characterized by notable changes in brain structure and function ^6^. Early by the age of seven months, infant language showed an inverse correlation with lower SES background ^7^. Also, adolescents from lower SES background have less access to enriching opportunities and resources, which could shape how adolescents respond to or pursue rewarding experiences ^8^. In the elderly, the relationship between SES and brain is complicated due to ‘cognitive reserve’, the ability to maintain cognitive abilities despite pathological changes ^9^. Several SES scales have been developed over the years, each using different indicators to measure SES. However, there are several common components used to incorporate SES scale; education level, occupation, income, geographic location, and self-reported of SES ^10, 11^.

Therefore, to address the effects of family income, and education level on brain glucose metabolism, we analyzed a large cohort of healthy middle-aged adults who underwent brain ^18^F- Fluorodeoxyglucose (FDG) positron emission tomography (PET), and survey of family income and education level. Human brain utilizes glucose as its main source of energy, thus, brain glucose metabolism, assessed by PET with ^18^F-FDG could be utilized for quantifying neuronal activity in the human brain ^12^. We used Bayesian hierarchical modeling to estimate the effects of family income, and education level on brain glucose metabolism and hypothesized that family income, and education level is positively associated with brain glucose metabolism.

## MATERIALS AND METHODS

### Subjects

We retrospectively analyzed data from 473 healthy males who underwent health check-up program at Samsung Changwon Hospital Health Promotion Center in 2013. After excluding subjects with neuropsychiatric disorders (n=5) or malignancies (n=3), those with missing data of family income or education level (n=232), 233 subjects were included in this study. Health check-up program included 1) Brain ^18^F-FDG PET, 2) anthropometric measurements, 3) survey of family income and education level, and 4) measures of stress, anxiety, and depression. Subjects in this study were included in a previous study of the effect of aging on brain glucose metabolism ^13^. The study protocol was approved by the Institutional Review Board of Changwon Samsung Hospital. The requirement for informed consent was waived owing to the retrospective study design.

### Brain ^18^F-FDG PET and Image analysis

Subjects were asked to avoid strenuous exercise for 24 hours and fast for at least 6 hours before PET study. PET/CT was performed 60 mins after injection of ^18^F-FDG (3.7 MBq/kg) with Discovery 710 PET/CT scanner (GE Healthcare, Waukesha, WI, USA). Continuous spiral CT was obtained with a tube voltage of 120kVp and tube current of 30-180mAs. PET scan was obtained in 3-dimensional mode with full width at half maximum of 5.6 mm and reconstructed using an ordered-subset expectation maximization algorithm. PET scans were spatially normalized to MNI space using PET templates from SPM5 (University College of London, UK) with pmod version 3.6 (PMOD Technologies LLC, Zurich, Switzerland). Automated Anatomical Labeling 2 (AAL2) atlas ^14^ was used to define region-of-interests (ROIs). The mean uptake of each ROI was scaled to the mean of global cortical uptake of each individual, and defined as standardized uptake value ratio (SUVR). For a full-volume analysis, the statistical threshold was set at a cluster level and corrected with false discovery rate with p < 0.05 in a regression model (correction with age) after smoothing SUVR images with a Gaussian kernel of FWHM 8mm (Statistical Parametric Mapping 12, Wellcome Centre for Human Neuroimaging, UCL, London, UK).

### Family income and Education level

Self-reported family income for the last 12 months (Korean won, KRW) was converted to US dollars (USD) with an exchange rate of 1,388 KRW. Education level was defined as the highest level of education completed by each subject; completion of 1) elementary school (6 years), 2) middle school (9 years), 3) high school (12 years), 4) 2-year college (14 years), 5) 4-year college (16 years), 6) graduate school (18 years).

### Measures of stress, anxiety, and depression

Stress measures for Korean National Health and Nutrition Examination Survey (KNHANES) consists of 9 self-reported items with higher scores indicating more stress ranging from 9 to 45 ^15^. Beck Anxiety Inventory is a 21 self-report measures with higher scores indicating more anxiety ranging from 0 to 63 ^16^. Centre for Epidemiologic Studies Depression Scale (CES-D) consists of 20 self-reported items with higher scores indication more depressed ranging from 0 to 60 ^17^.

### Neurosynth Image Decoder

To test the association of family income dependent brain glucose metabolism with Neurosynth terms, we used Neurosynth Image Decoder (https://neurosynth.org/decode). The t value map of the association between brain glucose metabolism and family income was uploaded on Neurosynth Image Decoder, and four meta-analytic uniformity maps of with the most similarity were downloaded.

### Statistical analysis

Spearman correlation was used to determine the association of family income with education level, and measures of stress, anxiety, and depression. We standardized the continuous variables of age, education level, family income, and log-transformed regional SUVR. Mediation analysis was performed, where each potential mediator (stress, anxiety, and depression) was tested separately in each model with family income or education level as a predictor and regional SUVR as the outcome. The effects of family income and education level on regional SUVR were investigated using Bayesian hierarchical modelling with brms ^18–20^ that applies the Markov-Chain Monte Carlo sampling tools of RStan ^21^. We set up a model for family income and education level with regional SUVR as a dependent variable and family income and education level as predictors adjusting for age. These fixed effects (family income, education level, and age) were calculated individually and subject and ROI were added as random intercepts to allow SUVR to vary between subjects and ROIs. Bayesian models were estimated using four Markov chains, each of which had 4,000 iterations including 1,000 warm-ups, thus totaling 12,000 post-warmup samples. The sampling parameters were slightly modified to facilitate convergence (max treedepth = 20). Total, direct, and indirect effects were calculated. Statistical analysis was carried out in R Statistical Software ver 4.4.1 (The R Foundation for Statistical Computing).

## RESULTS

### Subjects’ characteristics

A total of 233 healthy males (mean age: 42.6±3.5 years) were included in this study. The distribution of regional SUVR is shown in Figure 1. The mean family income was 61,319±17,978 USD; 32 had less than 50,000 USD, 193 between 50,000 USD and 100,000 USD, 8 more than 100,000 USD. The mean education level was 13.6±2.1 years; 1 completed elementary school; 133, high school; 28, 2-year college; 58, 4-year college; 13, graduate school. The subjects’ characteristics are summarized in Table 1. Family income was positively correlated with education level (rho=0.2580; p<0.0001), while the measures of stress (rho=0.0084; p=0.8979), anxiety (rho=-0.1224; p=0.0620), and depression (rho=-0.0338; p=0.6070) did not show the significant association with family income (Figure 1).

**Figure 1.**
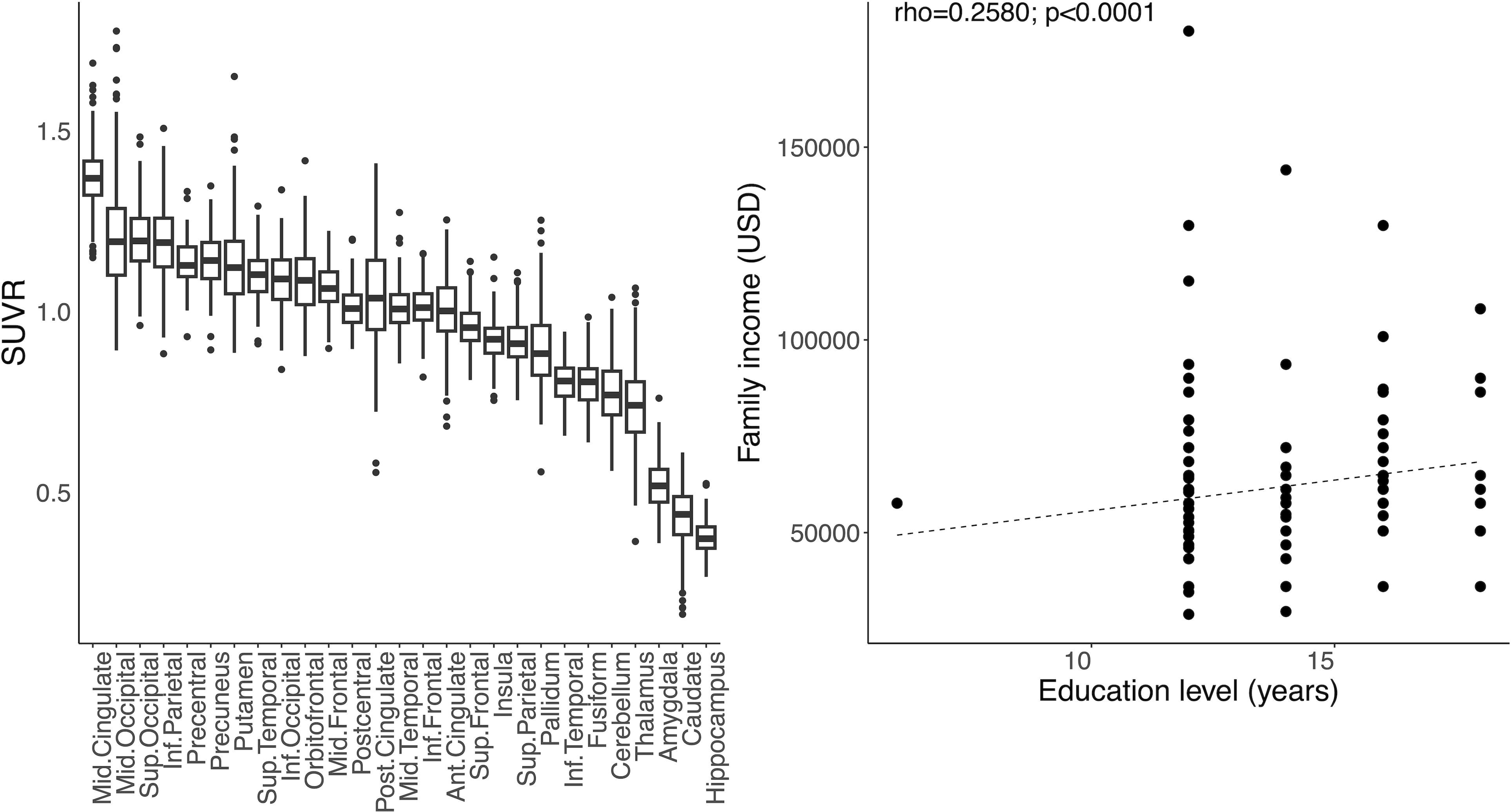
The distribution of regional SUVR and the association between family income (USD) and education level (years).

### Mediation of the association between family income, education level, and Brain glucose metabolism

The direct effect of family income on brain glucose metabolism was significant in amygdala, hippocampus, pallidum, putamen (p<0.05), however, that of education level was not significant in any regional brain glucose metabolism. There was no significant indirect effect of family income or education level via stress, anxiety, or depression on regional brain glucose metabolism.

### Family income, education level and Brain glucose metabolism

In a Bayesian model, family income was associated with brain glucose metabolism of fusiform, putamen, thalamus, anterior cingulate, pallidum, amygdala, hippocampus, and caudate showing some of their 95% posterior intervals overlapping with zero. However, educational level was not associated with regional brain glucose metabolism (Figure 2). Full-volume analysis revealed the consistent finding that shows the positive association of family income with brain glucose metabolism of caudate, putamen, hippocampus, amygdala, and anterior cingulate (Figure 3).

**Figure 2.**
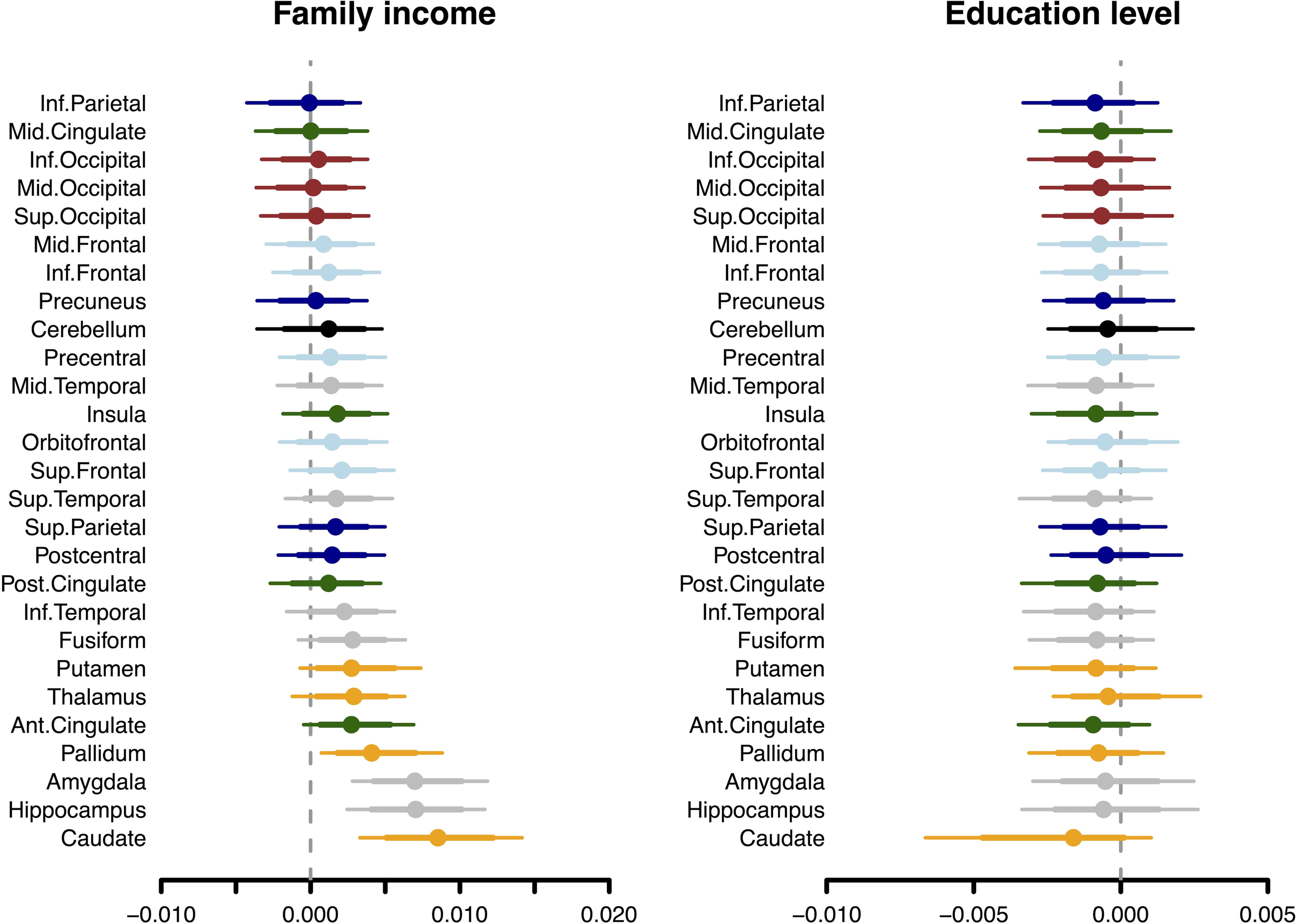
Posterior intervals of the regression coefficients for family income and education level predicting brain glucose metabolism. The thick lines represent the 80% posterior intervals, the thin lines represent the 95% posterior intervals, and the circles represent posterior means.

**Figure 3.**
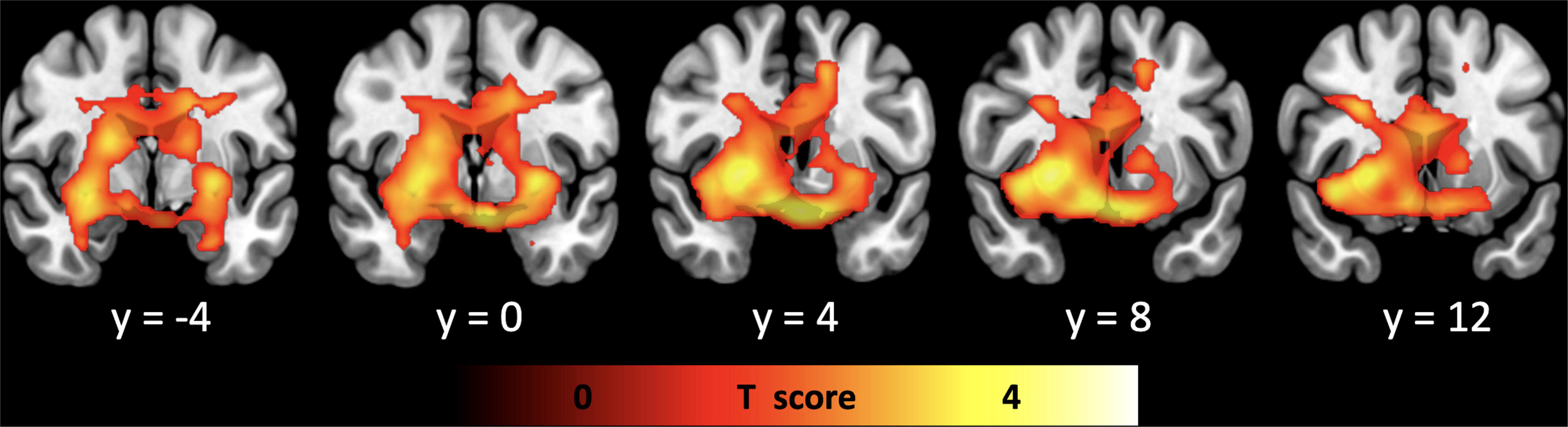
Full volume analysis showing the positive association of family income with brain glucose metabolism

### Localization of family income-dependent brain glucose metabolism and Neurosynth terms

Meta-analytic uniformity maps of reward (correlation coefficient 0.251), monetary (correlation coefficient 0.204), incentive (correlation coefficient 0.198), and motivation (correlation coefficient 0.182) showed the most similarity with family-income dependent brain glucose metabolism. The spatial correlation between family income-dependent brain glucose metabolism and four meta-analytic uniformity maps showed the significant similarity (Figure 4).

**Figure 4.**
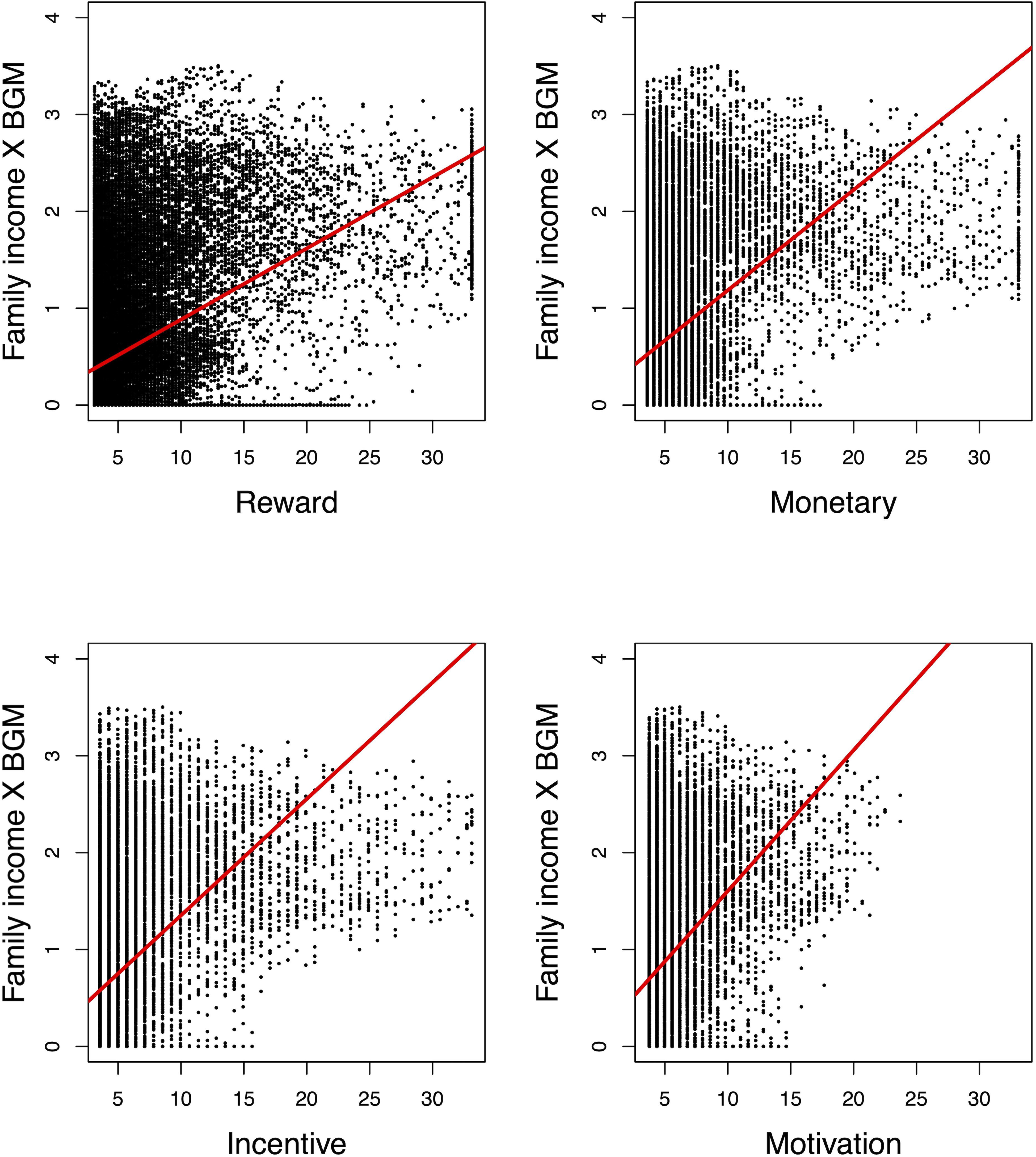
Spatial correlation between family income-dependent brain glucose metabolism and meta-analytic blood oxygenation level-dependent functional MRI activation patterns for four terms with the most similarity retrieved from the Neurosynth Image Decoder.

## DISCUSSION

Our main finding is that family income is positively associated with brain glucose metabolism in caudate, putamen, anterior cingulate, hippocampus, and amygdala, while education level does not show any significant association with brain glucose metabolism in middle-aged adults.

No human brain exists outside of a particular socioeconomic context ^2^. SES is an important factor relating to health and behaviors in mammals, including humans ^3^. SES also defines our social standing and has a significant impact on how our environment is shaped ^3^. Several SES scales have been developed over the years, each using different indicators to measure SES. The Hollingshead scale provides a composite score of social status based on the subject’s education level, and occupation ^10^. The Barratt Simplified Measure of Social Status (BSMSS) scale incorporates education level, and occupation of the subject, subject’s parents, and spouse, recognizing that social status is partly determined by the opportunities provided by one’s background ^11^. SES predicts a wide range of significant life outcomes. Higher SES is associated with better physical health; incidences of cancer, heart disease, stroke, diabetes, psychosis, depression, and anxiety decline with SES, and lifespan is positively correlated with SES ^4^. However, SES is often included in neuroimaging studies as a ‘nuisance variable’, covariates of no interest ^22^. Also, examination of the relationship between SES, and the brain has primarily focused on the earlier or later stages of the lifespan when the brain is most vulnerable, which are characterized by notable changes in brain structure and function ^6^. In addition, SES variables such as education level, family income, or occupation may not have a consistent association with brain across subjects with varying ages and social contexts ^23^. Therefore, we evaluated the association of the elements of SES (family income, and education level) with regional brain glucose metabolism in middle-aged adults.

In a study of social dominance in monkeys, dominant monkeys had higher DR availability compared to subordinate monkeys after three months of social housing ^5^. Comparing with the baseline state of the individual housing, DR availability was increased only in dominant monkeys, not in subordinate monkeys after the social structure was established ^5^. In addition, subordinate monkeys were more susceptible to the reinforcing effects of cocaine ^5^. Also, in humans, SES scores measured with the Hollingshead scale ^24^ or BSMSS ^25^ were positively correlated with DR availability in the striatum, suggesting that the higher social status, a greater sense of perceived social support, and lower levels of social avoidance are associated with higher DR availability ^25^. Therefore, social context can have profound effects on brain dopaminergic function in both nonhuman primates and humans. In addition, according to a recent study, adolescents with higher SES background showed greater reward-driven activation than those with lower SES background in the caudate, putamen, suggesting that lower SES environments reduce reward sensitivity ^26^. Also, in adults with lower SES background, the response to negatively valenced stimuli was higher from cortex, and the response to rewarding stimuli was lower from anterior cingulate, striatum than those with higher SES background, probably due to the altered functional connectivity implicated in reward processing and impulse regulation ^27^. Considering that most previous studies analyzed the effect of SES of parents in the childhood, this study investigated the association between family income and brain glucose metabolism more directly, as all subjects included in this study are employed adults who underwent health check-up program as corporate welfare. Therefore, the strong link between family income and glucose metabolism of caudate, putamen, and anterior cingulate in this study might reflect the neuronal activation of reward system, interpreted as the increased reward sensitivity in higher family income, which is consistent with the findings from Neurosynth Image Decoder.

In adults who reported financial hardship, the volume of hippocampus and amygdala was smaller than those who did not report financial hardship. Financial hardship is a potent stressor, and might affect the volume of hippocampus and amygdala via the action of the hypothalamic-pituitary-adrenal axis and other stress-related pathways ^28^. Also, equivalent income was associated with the volume of hippocampus-amygdala complex ^29^, reflecting the impact of chronic stress derived from disadvantageous life conditions ^30^. Similar with previous studies, family income was positively associated with brain glucose metabolism in hippocampus, and amygdala, however, no significant indirect effect of family income or education level via stress, anxiety, or depression was observed in this study.

Surprisingly, there was no brain region showing the significant association with education level both from ROI analysis and full volume analysis in this study, although family income and education level was significantly correlated. According to previous studies, education level was positively correlated with brain glucose metabolism ^9^ and gray matter volume ^29^ of anterior cingulate. The cognitive reserve hypothesis posits that brains with higher reserve could tolerate more aging or pathological effects, thereby minimizing the symptoms in elders with higher education level ^9^. However, the subjects included in previous studies (mean age of 67.1, 52.4 years) are older than this study (mean age 42.6 years). Therefore, the effect of education level on the brain might not be substantial until the age of 40s before the aging process accelerates.

This study has several limitations. Firstly, only males were included in this study, thus these results may not directly generalize to females. Secondly, this retrospective study was based on health check-up program. Therefore, a brain MRI was not included in the program, and MRI-based coregistration and partial volume correction of PET scans could not be done. Thirdly, self-reported family income for the last 12 months was included in this study, however, there are several other measures of financial status such as income-to-needs ratio, or equivalent income. Lastly, it is unclear whether this observed association between family income and brain glucose metabolism reflects a neurobiological predisposition, or a neurobiological alteration induced by the attainment of income.

In conclusion, family income, and education level show heterogenous relationships with brain glucose metabolism. Family income is positively associated with brain glucose metabolism in caudate, putamen, anterior cingulate, hippocampus, and amygdala, while education level does not show any significant association with brain glucose metabolism. This finding might reflect the link between family income, and reward sensitivity, stress in middle-aged adults.

## Supporting information

tablle

## Data Availability

All data produced in the present study are available upon reasonable request to the authors

## ACKNOWLEDGEMENTS

No

## CONFLICT OF INTEREST

The authors declare no conflict of interest

## Notes

### Competing Interest Statement

The authors have declared no competing interest.

### Funding Statement

This study did not receive any funding

### Author Declarations

The study protocol was approved by the Institutional Review Board of Changwon Samsung Hospital.

