## Supplementary material for "Family income is associated with regional brain glucose metabolism in middle-aged adults": tablle

**TABLES**

Table 1. Subjects’ characteristics

|  | Mean±Standard deviation |
| --- | --- |
| Age (years) | 42.6±3.5 |
| Height (m^2^) | 1.7±0.1 |
| Weight (kg) | 73.2±9.9 |
| Body mass index (kg/m^2^) | 24.6±2.9 |
| Waist-hip ratio | 9.7±3.3 |
| Measures  Stress  Anxiety  Depression | 16.1±6.2  4.9±6.0  7.6±5.9 |
| Family income (USD) | 61319±17978 |
| Education level (years) | 13.6±2.1 |
